# Assisted reproductive technology and association with childhood cancer subtypes

**DOI:** 10.1101/2021.04.26.21256117

**Authors:** Natalie B. Gulrajani, Samuel Montes, Dan McGough, Courtney E. Wimberly, Ameera Khattab, Eleanor C. Semmes, Lisa Towry, Jennifer L. Cohen, Jillian H. Hurst, Daniel Landi, Sherika N. Hill, Kyle M. Walsh

**Author notes:** **Address correspondence to:** Kyle M. Walsh, DUMC Box 3050, Durham, NC 27710. **Conflict of Interest Disclosures:** The authors have no conflicts of interest to disclose. **Funding/Support:** This study was supported by a grant from the Alex’s Lemonade Stand Foundation (KMW). **Role of Funder/Sponsor:** This work was supported by a grant from the Alex’s Lemonade Stand Foundation (ALSF). The funder hosts and advertises the survey portal from which data in this manuscript were collected. The funder had no role in data analysis or interpretation, although an employee of ALSF (L.T.) is a co-author of this manuscript in recognition of her significant contributions to constructing and piloting the surveys and her participation in reviewing the final version of the manuscript presented here.

## Abstract

**Objectives:** Use of assisted reproductive technology (ART) may alter the typical course of fetal development. We sought to investigate the association between ART use and childhood cancer subtype.

**Study design:** We surveyed 1701 parents of children with cancer about ART use, demographics, and gestational and perinatal factors. Multivariable logistic regression modeled the association between ART use and childhood cancer subtypes, birthweight and multiple gestation status.

**Results:** Among childhood cancer patients, ART use was highest among children with osteosarcoma (OR=2.81; 95% CI=1.2-6.4; P=0.01). ART use was also elevated among children with hepatoblastoma, and this relationship appeared mediated by low birthweight. No specific type of ART appeared to drive these associations. Low birthweight was itself strongly associated with increased hepatoblastoma risk, even after adjustment for ART use, multiple gestation status, sex, and parental income (P<0.001). Birthweight was higher in patients with germ cell tumors (P=0.02) and with neuroblastoma (P=0.06). Multiple gestation status was associated with neuroblastoma among females (OR=3.6, 95% CI=1.2-10.5, P=0.02), but not among males (OR=0.97, 95% CI=0.27-3.4, P=0.96) (P_interaction_=0.02).

**Conclusions:** Associations between ART use and hepatoblastoma risk may be mediated by birthweight, a strong hepatoblastoma risk factor that was replicated in our study. ART use may also be associated with osteosarcoma independent of birthweight, an association not previously observed in studies limited to cancers diagnosed before adolescence. Evaluating long-term health outcomes in children conceived by ART, throughout adolescence and potentially into adulthood, appears warranted.

## INTRODUCTION

Cancer is a leading cause of death in children ages 5-14 years.^1^ While mortality rates have decreased and 5-year survival rates exceed 80%, nearly 2000 childhood cancer-associated deaths occur annually in the U.S.^2^ Epidemiologic data suggest a modest increase in childhood cancer incidence, although the factors underlying this trend remain largely unresolved.^3^ One possibility is that the prevalence of underlying risk factors may be changing. In the U.S., the incidence of acute lymphoblastic leukemia (ALL) is highest in the Hispanic population, and as this population has grown, national rates of ALL have risen accordingly.^4^ Whether increased cancer incidence is also driven by modifiable risk factors merits further evaluation.

While childhood cancer incidence has increased overall, this pattern does not hold across all subtypes. For example, rates of hepatoblastoma and central nervous system (CNS) tumors increased substantially from 1992-2004.^3^ The increase in CNS tumor incidence is partially attributable to incidental detection of indolent tumors with the expanded use of medical imaging;^5^ however, factors underlying increases in hepatoblastoma incidence remain unclear.

Additionally, trends in childhood cancer incidence differ by race and ethnicity, suggesting non-uniform changes in the prevalence of underlying risk factors across populations.^6^

Although numerous genetic factors increase childhood cancer risk, relatively few modifiable risk factors have been conclusively identified. Several pre- and perinatal factors have been associated with childhood cancer risk, including *in utero* exposure to ionizing radiation,^7^ very high or very low birthweight,^8^ *in utero* diethylstilbestrol exposure,^9^ and congenital cytomegalovirus (CMV) infection.^10,11^ Such findings suggest that small perturbations to cellular differentiation during gestation may have outsized effects on both normal and malignant cellular development.

The use of assisted reproductive technologies (ART) to conceive is increasingly common and has previously been associated with elevated childhood cancer risk.^12–14^ Because ART is a broad term that includes many types of infertility treatment, including in-vitro fertilization (IVF), use of donor sperm or eggs, intracytoplasmic sperm injection (ICSI), gamete intrafallopian transplantation (GIFT), intrauterine insemination (IUI), frozen embryo transfer (FET), and fertility drugs, each ART method could have substantially different impacts on *in utero* development. ART use has been associated with reduced birthweight, increased birthweight (in the case of FET), multiple gestations, a sex-ratio skewed toward male births, and increased occurrence of congenital/developmental abnormalities.^15–19^ These observations indicate that ART may alter the typical course of fetal development, with potentially different effects across ART modalities.

To evaluate the association of ART with subtype-specific childhood cancer risk, we performed a case-only analysis of childhood cancer patients from the Alex’s Lemonade Stand Foundation’s *My Childhood Cancer: Survey Series* cohort. Parents representing >1,700 childhood cancer patients provided data on ART utilization, multiple gestation status, birthweight, and other demographic and pre/perinatal factors. We sought to understand the potential role of ART in contributing to subtype-specific childhood cancer risk, accounting for contributions from other prenatal/perinatal factors.

## METHODS

### Study Population

To explore associations between ART use and childhood cancers, we partnered with the Alex’s Lemonade Stand Foundation (ALSF) to conduct an ongoing series of longitudinal surveys of families affected by childhood cancer. Initiated in 2011, the ALSF *My Childhood Cancer* (*MCC*): *Survey Series* explores families’ experiences and attitudes from diagnosis, throughout treatment and follow-up care, and after bereavement (when applicable).^20^ To date, 3150 families have participated in the *MCC* survey series. In this study, we examined responses to the ALSF MCC diagnosis survey completed between August 2012 and April 2019, as prior versions of the survey did not collect data on ART use. Thus, analyses for this study were performed on the subgroup with available ART data (N=1701 respondents), limiting to one parental respondent per family. Median time from diagnosis to survey completion was 2 years.

### Survey Instruments

Childhood cancer type and patient/parental demographics are collected at *MCC: Survey Series* registration. Survey respondents were asked “Did [child]’s biological mother receive any medical help in order to become pregnant with [child]?” Responses of “Yes” and “No” were collapsed into an indicator for ART use. Those who responded “Not sure” were excluded. Those who indicated that they received medical help to conceive were asked to answer whether any or all of the following methods of ART were used: IVF, fertility drugs, donor sperm/donor eggs, ICSI, IUI, and GIFT. At the time of data analysis, FET was not a specified ART modality in the survey, but has since been added. We assume that any respondents whose child was conceived via FET would have answered “Yes” to the ART question, and may have additionally endrsed additional ART modalities (*e.g*., “fertility drugs”). The “Yes” and “No” answers to these questions along with the “No” answers to any ART use were collapsed into binary indicators for each ART type. Respondents who reported use of more than one ART type were included in each specific ART type variable used, but were counted only once in the overall ART use variable. Respondents recorded child’s birthweight in categories to the nearest pound in the following bins: “3 pounds or less”, “4-5 pounds”, “6-9 pounds”, “10-11 pounds”, “12 pounds or more”, and “Not sure”. Birthweight was modeled as an ordinal variable. A question about whether the child with cancer was a singleton or multiple gestation was collapsed into an indicator for multiparity. Respondent race/ethnicity was collapsed into an indicator for “non-Hispanic white” versus “American Indian/Alaskan Native,” “Native Hawaiian/Pacific Islander,” “Black/African American,” “Other race,” and “Hispanic or Latino (of any race).” Household income was recorded in the following bins: <$20,000; $20,000-$49,999; $50,000-$74,999; $75,000-$99,999; $100,000-$149,999; ≥$150,000 and modeled as an ordinal variable. Median time from diagnosis to survey completion was 2 years and did not differ across cancer subtype.

Dependent variables were specific childhood cancer subtype, including: Hodgkin’s lymphoma, non-Hodgkin’s lymphoma, germ cell tumor, Kidney/Wilms’ tumor, hepatoblastoma/liver cancer, neuroblastoma, retinoblastoma, rhabdomyosarcoma, osteosarcoma, Ewing sarcoma, an “all sarcomas” subgroup, ALL, and acute myeloid leukemia (AML). We also included three brain/spinal tumor subgroups: primitive neuroectodermal tumors (PNETs) including medulloblastoma, supratentorial PNETs, and atypical teratoid/rhabdoid tumor (AT/RT); glioma, including astrocytoma/anaplastic astrocytoma, juvenile pilocytic astrocytoma (JPA), diffuse intrinsic pontine glioma (DIPG), glioblastoma; and ependymoma.

### Statistical Analyses

Independent variables of interest in this analysis included ART use, ART type, birthweight, and multiple gestation status. Survey respondent’s race/ethnicity, household income, and years between child’s birth and survey completion were included in models as potential confounders (as parents of children diagnosed at older ages would be reporting perinatal factors from longer ago, which could result in differential exposure misclassification). We also included child’s sex in models because it could potentially act as a confounder (*e.g*., boys generally have higher birthweight than girls and male children are at increased risk of CNS tumors)^21^ or as an effect modifier (which was evaluated using interaction terms).

Relationships between independent and dependent variables were assessed using chi-square tests for independence or t-tests for difference. Fisher’s exact tests were used when analyzing specific ART types due to small sample sizes. For multivariate analyses, logistic regression was used to examine potential relationships between variables. For all statistical tests, alpha = 0.05 was used to determine statistical significance. Stata version 16.0 was used for data analysis.

## RESULTS

Between August 2012 and April 2019, a total of 1701 respondents from unique families completed the diagnosis survey asking about use of ART to conceive. Respondents were largely female, the biologic parent, and identified as non-Hispanic white. The majority of children were 6-9 pounds at birth, singletons, and were only-children at the time of diagnosis (**Table 1**). 126 respondents (7.4%) reported using ART to become pregnant, of which 85% specified the type of ART and 36% reported using more than one form of ART. Parental ART use was most frequent among children with osteosarcoma (16%), followed by hepatoblastoma (14%), and was least common among children with germ cell tumors (4%) and retinoblastoma (0%) (**Figure 1**).

**Table 1.** Demographic characteristics of survey respondents

| **Table 1.** Demographic characteristics of survey respondents |  |  |
| --- | --- | --- |
|  | **Number of**<br>**respondents**<br>**(n=1701)** | **Proportion of**<br>**study population**<br>**or mean (SD)** |
| **Respondent biologic parent** | 1673 | 0.98 |
| **Respondent gender** |  |  |
| Female | 1589 | 0.93 |
| Male | 111 | 0.07 |
| Missing | 1 | <0.01 |
| **Respondent race/ethnicity** |  |  |
| Non-Hispanic white | 1582 | 0.93 |
| Other | 105 | 0.06 |
| Missing | 14 | 0.01 |
| **Birthweight** |  |  |
| 3 pounds or fewer | 26 | 0.02 |
| 4-5 pounds | 107 | 0.06 |
| 6-9 pounds | 1468 | 0.86 |
| 10-11 pounds | 92 | 0.05 |
| 12 pounds or more | 5 | <0.01 |
| Missing | 3 | <0.01 |
| **Household income at**<br>**diagnosis** |  |  |
| Less than \$20,000 | 129 | 0.08 |
| \$20,000-\$49,999 | 368 | 0.22 |
| \$50,000-\$74,999 | 365 | 0.21 |
| \$75,000-\$99,999 | 313 | 0.18 |
| \$100,000-\$149,999 | 254 | 0.15 |
| \$150,000+ | 180 | 0.11 |
| Missing | 92 | 0.05 |
| **Child Sex** |  |  |
| Female | 933 | 0.55 |
| Male | 767 | 0.45 |
| Missing | 1 | <0.01 |
| **ART Use<sup>a</sup>** |  |  |
| Any ART | 126 | 0.07 |
| IVF | 27 | 0.02 |
| Fertility drugs | 103 | 0.06 |
| Donor Sperm/Eggs | 10 | 0.01 |
| ICSI | 13 | 0.01 |
| IUI | 24 | 0.01 |
| GIFT | 1 | <0.01 |
| Missing | 18 | 0.01 |

| **Table 1.** Continued |  |  |
| --- | --- | --- |
| **Child age at diagnosis** | ... | 6.25 (4.97) |
| **Siblings** |  |  |
| None | 1301 | 0.76 |
| 1+ | 384 | 0.23 |
| Missing | 16 | 0.01 |
| **Multiparity** |  |  |
| Singleton | 1625 | 0.96 |
| Multiple gestation | 68 | 0.04 |
| Missing | 8 | <0.01 |
| Abbreviations: ART, assisted reproductive technology; IVF, *in vitro* fertilization; ICSI, intracytoplasmic sperm injection; IUI, intrauterine insemination; GIFT, gamete intrafallopian transfer. |  |  |
| <sup>a</sup>Respondents could choose multiple types of ART, if applicable. |  |  |

**Figure 1:**
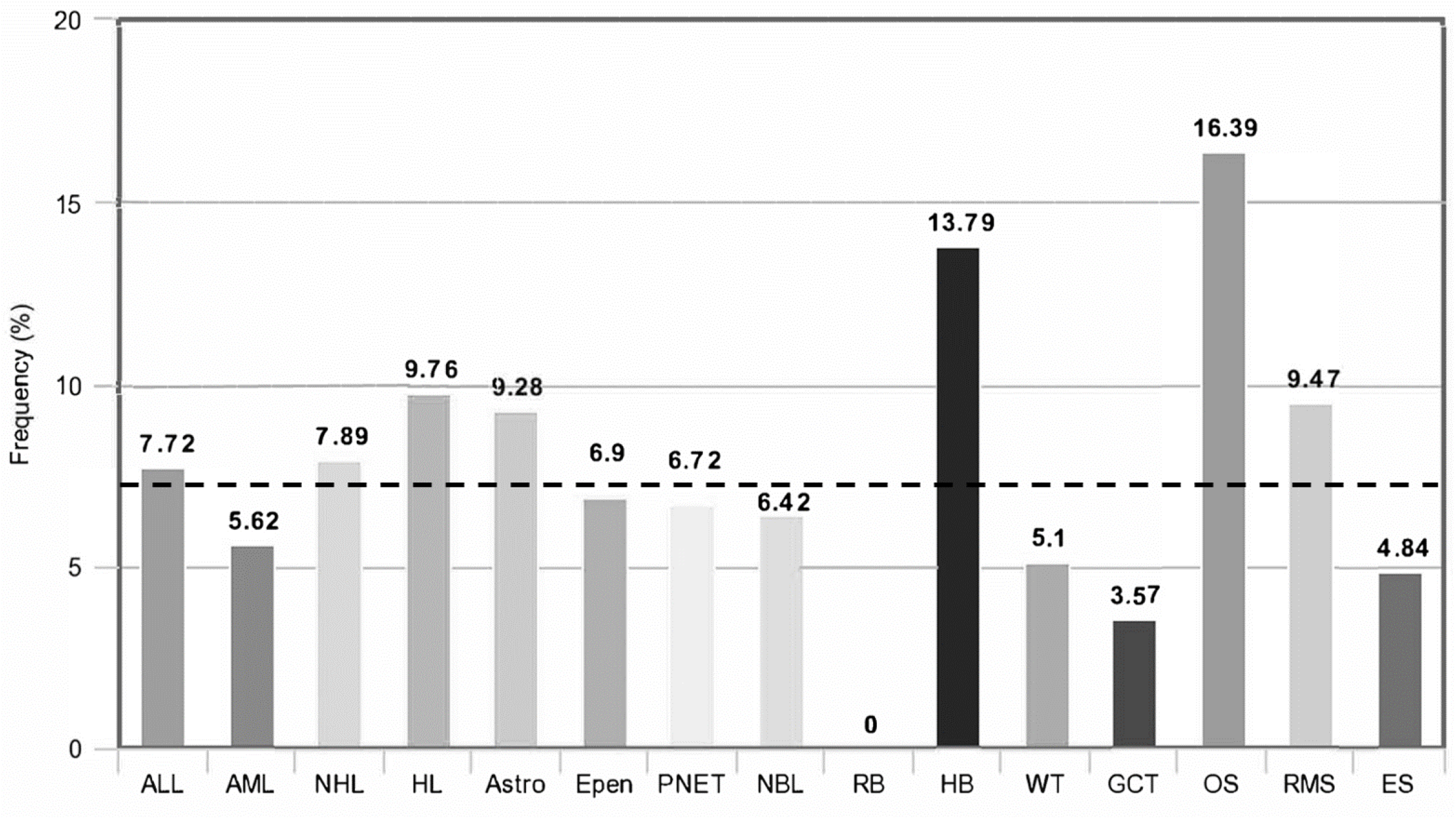
Proportion of children in the MCC cohort, stratified by cancer type, who were conceived with use of Assisted Reproductive Technology. The dashed horizontal line indicates the rate of ART usage across all cancer types, combined (7.4%). Cancer types, from left to right, are: Acute lymphoblastic leukemia (ALL, N=492), Acute myeloid leukemia (AML, N=89), non-Hodgkin lymphoma (NHL, N=76), Hodgkin lymphoma (HL, N=41), Astrocytoma (Astro, N=97), Ependymoma (Epen, N=29), primitive neuroectodermal tumors including medulloblastoma, AT/RT and supratentorial PNETs (PNET, N=119), Neuroblastoma (NBL, N=187), Retinoblastoma (RB, N=28), Hepatoblastoma (HB, N=29), Wilms tumor (WT, N=98), Germ cell tumors (GCT, N=28), Osteosarcoma (OS, N=55), Rhabdomyosarcoma (RMS, N=95), Ewing sarcoma (ES, N=62).

Univariate relationships between cancer subtype and ART use, multiparity, and birthweight were tested (**Table S1**). ART use was associated with a 2.6-fold increased odds of osteosarcoma relative to all other cancer types (OR=2.56; 95% CI=1.22-5.35; P=0.01) and a 2-fold increased odds of hepatoblastoma (OR=2.03; 95% CI=0.70-5.93; P=0.19), although this latter association did not reach statistical significance. Multiple gestation was associated with a 2.7-fold increased odds of hepatoblastoma (OR=2.71; 95% CI=1.14-6.42; P=0.02) and a 1.6-fold increased odds of neuroblastoma (OR=1.62; 95% CI=1.03-2.54; P=0.03). Children with hepatoblastoma had significantly lower birthweight than children with other cancer types (P<0.001), while children with germ cell tumors had modestly higher birthweight than children with other cancer types (P=0.08) (**Table S1**).

In the entire sample, ART use was associated with lower birthweight (p<0.001) and with being a multiple gestation (p<0.001). Multiple gestations were also associated with lower birthweight (p<0.001). To disentangle these correlated effects, we modeled the association of each childhood cancer subtype with ART use, multiple gestation status, birthweight, and child sex in a multivariable model that adjusted for race/ethnicity, household income, and years between birth and survey completion. In this multivariable model, the association between ART use and osteosarcoma persisted (OR=2.81; 95% CI=1.24-6.35; P=0.01). For hepatoblastoma, however, the magnitude of effect from univariate analyses (OR=2.03) was strongly attenuated in the multivariable model (OR=1.39; 95% CI=0.41-4.70; P=0.60) (**Table 2**). Univariate associations between multiple gestation status and both hepatoblastoma and neuroblastoma were also strongly attenuated in multivariable models. Low birthweight, however, remained strongly associated with hepatoblastoma in the multivariable model (P<0.001). The association between higher birthweight and germ cell tumor risk was strengthened in the multivariable model, reaching statistical significance (P=0.02) (**Table 2**).

**Table 2.** Relationships between ART use, multiparous birth, birthweight and cancer type^a^

| Cancer Type | ART use<br>(OR<sup>c</sup>, 95% CI) | P-value | Multiparous birth<br>(OR<sup>c</sup>, 95% CI) | P-value | Birthweight<sup>b</sup><br>(OR<sup>c</sup>, 95% CI) | P-value |
| --- | --- | --- | --- | --- | --- | --- |
| Hodgkin lymphoma (N=41) | 1.22 | 0.73 | 1.75 | 0.42 | 0.92 | 0.82 |
| Non-Hodgkin lymphoma (N=76) | 1.23 | 0.64 | 0.61 | 0.53 | 0.96 | 0.89 |
| Germ cell tumor (N=28) | 0.54 | 0.55 | NA | NA | **2.70** | **0.02** |
| Wilm's tumor (N=98) | 0.71 | 0.48 | 1.22 | 0.73 | 0.97 | 0.91 |
| Hepatoblastoma (N=29) | 1.39 | 0.60 | 1.62 | 0.48 | **0.35** | **<0.001** |
| Neuroblastoma (N=187) | 0.76 | 0.42 | 1.87 | 0.12 | 1.45 | 0.058 |
| Retinoblastoma (N=28) | NA | NA | NA | NA | 0.65 | 0.32 |
| Rhabdomyosarcoma (N=95) | 1.22 | 0.62 | 1.17 | 0.78 | 1.16 | 0.57 |
| Osteosarcoma (N=55) | **2.81** | **0.01** | 0.73 | 0.69 | 1.09 | 0.79 |
| Ewing sarcoma (N=62) | 0.64 | 0.48 | 1.07 | 0.93 | 1.18 | 0.61 |
| All sarcomas (N=250) | 1.31 | 0.31 | 1.27 | 0.51 | 1.02 | 0.93 |
| ALL (N=492) | 1.16 | 0.73 | 0.73 | 0.61 | 1.16 | 0.56 |
| AML (N=89) | 0.75 | 0.58 | 0.52 | 0.43 | 0.70 | 0.21 |
| PNET<sup>d</sup>(N=119) | 0.95 | 0.91 | 0.79 | 0.71 | 1.23 | 0.40 |
| Ependymoma (N=29) | 1.04 | 0.96 | 0.90 | 0.92 | 0.98 | 0.97 |
| Astrocytoma<sup>e</sup> (N=97) | 1.17 | 0.70 | 1.84 | 0.24 | 1.59 | 0.07 |

Abbreviations: ART, assisted reproductive technology; PNET, primitive neuroectodermal tumor; AT/RT, atypical teratoid/rhabdoid tumor; DIPG, diffuse intrinsic pontine glioma.
<sup>a</sup>Multivariable logistic regression, including child sex, household income, respondent race/ethnicity.
<sup>b</sup>Birthweight was recorded in the following categories and modeled as an ordinal variable: ≤3 pounds, 4-5 pounds, 6-9 pounds, 10-11 pounds, ≥12 pounds
<sup>c</sup>Odds ratios are from case-case analyses, comparing the listed cancer subtype to all other childhood cancer patients in the study. Therefore, it should be noted that the reference group changes slightly for each subtype-specific comparison.
<sup>d</sup>Including medulloblastoma and AT/RT
<sup>e</sup>Including pilocytic, glioblastoma, and DIPG

Inclusion of a “sex*ART” or “sex*birthweight” interaction term generally did not improve multivariable model fit for osteosarcoma, hepatoblastoma, neuroblastoma, or germ cell tumors. Inclusion of a “sex*multiple gestation status” term in the neuroblastoma model indicated that multiple gestation status was significantly associated with neuroblastoma in females (OR=3.59, 95% CI=1.23-10.50, P=0.02), but not in males (OR=0.97, 95% CI=0.27-3.41, P=0.96) (P_interaction_=0.02).

To isolate the effect of specific ART modality on cancer subtype, additional analyses were conducted for osteosarcoma and hepatoblastoma, which had the highest univariate odds ratios with ART use (**Table S2**). Every ART modality was similarly positively associated with hepatoblastoma and osteosarcoma, although no association reached statistical significance due to the limited sample size for each ART modality. Overall, our results did not identify any particular ART modality that appeared to drive the overall association of ART use with hepatoblastoma and osteosarcoma.

## DISCUSSION

ART is an increasingly common method to overcome infertility, but perinatal risks associated with its use continue to raise concerns. Associations between ART use and birthweight, a skewed sex-ratio, congenital/developmental abnormalities, and potential associations with childhood cancer risk suggest that ART may alter typical fetal development.^12–18^ We observed an association between ART use and childhood osteosarcoma in our case-only analyses, independent of both birthweight and multiple gestation status. We also observed modest evidence of association between ART use and hepatoblastoma, potentially mediated by the strong association between low birthweight and hepatoblastoma risk. Associations between higher birthweight and increased risk of neuroblastoma and germ cell tumors were also observed, aligned with previous registry-based reports.^8,22,23^

Although associations between ART and childhood cancer risk have been studied previously, subtype-stratified risk and risk beyond the first decade of life remain unclear.^24^ Williams, *et al*. conducted a population-based study of all children born in Britain from 1992-2008 and observed an increased risk of hepatoblastoma and rhabdomyosarcoma in association with ART use. The study assessed potential mediating and moderating factors, and attributed hepatoblastoma risk primarily to low birthweight in association with ART use.^25^ More recently, Spector, *et al*. observed that hepatoblastoma risk was significantly associated with IVF use, but they did not include birthweight in their modeling.^26^ Although they did not identify associations between ART use and osteosarcoma risk, the follow-up time after birth was approximately 8 years, making it unlikely that adolescent-onset cancers (*i.e*., osteosarcoma) would be captured. Although osteosarcoma risk is associated with taller adolescent and adult stature, recent registry studies have not observed an association between osteosarcoma risk and infant birth length or birthweight.^8,27,28^ Our analyses support the independence of osteosarcoma risk and birthweight, but implicate ART in osteosarcomagenesis, independent of sex, birthweight and multiple gestation status.

In addition to its association with birthweight and multiple gestations, ART use has also been associated with a sex-ratio skewed toward male births and an increased incidence of congenital/developmental abnormalities.^15–18^ A recent murine study identified impaired imprinting of the inactivated X chromosome as a primary epigenetic barrier for female embryo development that was responsible for sex-skewing toward male births following IVF.^29^ Such dysregulated epigenetic programming could lead to overexpression of key genes located in imprinted regions, both on the X chromosome and the autosomes, providing a potential molecular mechanism for these ART-associated phenotypes. Importantly, in murine models this epigenetic dysregulation was resolved through retinoic acid supplementation, providing a potential opportunity for risk interception.^29^

Similar to prior studies, we observed a two-fold increase in ART use among hepatoblastoma patients compared to children with other cancers. This association was not statistically significant given the modest number of hepatoblastoma patients in our dataset (N=61), and the magnitude of effect was greatly attenuated in multivariable models accounting for birthweight. Multiparity and low birthweight were significantly associated with hepatoblastoma risk in univariate analyses, but only the association with low birthweight persisted in multivariable models, suggesting that hepatoblastoma risk associated with multiparity or ART may be mediated by birthweight, corroborating previous studies.^8,30–32^

We observed that neuroblastoma patients were more likely to be part of a multiple gestation than children with other cancer types but did not identify an association between neuroblastoma and birthweight in univariate models. Multivariable models including both multiple gestation status and birthweight revealed a potential association between higher birthweight and neuroblastoma risk, while the effect of multiple gestation status remained fairly consistent. These results suggest that both higher birthweight and being the product of a multiple gestation may be associated with neuroblastoma, despite birthweight generally being lower in multiple gestations. That higher birthweight and increased twinning are paradoxically associated with increased neuroblastoma risk suggests that the effect of birthweight on neuroblastoma risk may be confounded toward the null in models that do not account for multiple gestation status. Interaction tests provide evidence that the effect of multiple gestation status on neuroblastoma risk may be female-specific, an observation that merits follow-up in future studies.

We also observed that patients with germ cell tumors had higher birthweight than children with other cancer types, consistent with prior reports.^8,22,23^ No other childhood cancer subtypes showed significant associations with ART use, multiple gestation status, or birthweight. Notably, we did not observe an association between ART use and rhabdomyosarcoma or ALL, both of which have been previously but not universally observed in prior studies.^24,25^

When looking at specific ART modalities in association with hepatoblastoma and osteosarcoma, each ART modality had similar positive magnitudes of effect; however, none reached statistical significance due to the small number of parents who utilized each ART moadlity. This analysis suggests that no single ART modality drove the overall associations with childhood cancer that were observed. Importantly, IVF was not the only modality with a large odds ratio, nor did it have the largest magnitude of effect across modalities, indicating a need for further studies on the association between ART use and childhood cancer etiology, especially studies that do not focus exclusively on IVF.

Our study has several limitations, including that survey participants were a self-selected population of caregivers who independently navigated to the ALSF MCC survey portal and do not represent a random sample of parents of children with cancer. A related limitation is that survey respondents are primarily non-Hispanic white and results may not be broadly generalizable; however, the distribution of household incomes in our sample aligns reasonably well with the income distribution of U.S. households. Although our study design does not permit a “disease-free” control group for case-control comparisons, our case-only analytic approach minimizes many of the biases common in case-control study designs, especially recall bias.

Parents of children diagnosed at older ages, such as osteosarcoma, may less accurately recall perinatal exposures from many years earlier, although adjusting for time between birth and survey completion did not meaningfully alter results. Our study is relatively large for an epidemiologic assessment of ART and childhood cancer, but several specific cancer types included in analyses were uncommon (*e.g*. 28 patients with retinoblastoma), limiting precision of our effect estimates. Another limitation to our study is potential confounding due to the unknown relationship between parental infertility itself and risk of cancer in children.

Our dataset of 1701 families affected by childhood cancer uses parent-reported data on ART use that do not rely on registry linkage. Birth registries often capture ART use unreliably and limit assessments to IVF, thus excluding other forms of ART.^33^ We assessed the effect of ART use on childhood cancer subtypes, including stratification by ART modality. Furthermore, because our study did not depend upon registry data, we were able to observe a potential association between ART use and risk of osteosarcoma, a malignancy frequently diagnosed too far into adolescence to be evaluated in studies relying on IVF databases with <10 years of follow-up.

Because ART is an increasingly prevalent means of conception, it is important to understand any potential associated risks. Our findings support prior investigations linking ART use to risk of hepatoblastoma, and suggest this association may be attributable to the mediating effect of low birthweight, an established hepatoblastoma risk factor. Finally, we observed a novel, birthweight-independent association between ART use and osteosarcoma risk, emphasizing the need to continue studying ART-associated cancer risks beyond the first ten years of life, into adolescence and perhaps continuing throughout adulthood.

## Data Availability

When paper is published, the de-identified, individual-level data are available from the authors upon reasonable request.

## Abbreviations

(ART): assisted reproductive technology
(ALSF): the Alex’s Lemonade Stand Foundation
(IVF): in vitro fertilization
(PCA): principal component analysis

